# The adverse impact of consecutive COVID-19 waves on mental health

**DOI:** 10.1101/2021.07.25.21261094

**Authors:** Jan Sebastian Novotný, Juan Pablo Gonzalez-Rivas, Šárka Kunzová, Mária Skladaná, Anna Pospíšilová, Anna Polcrová, Maria Vassilaki, Jose Ramon Medina-Inojosa, Francisco Lopez-Jimenez, Yonas Endale Geda, Gorazd Bernard Stokin

**Affiliations:** Translational Neuroscience and Aging Program, Centre for Translational Medicine, International Clinical Research Centre, St. Anne’s University Hospital in Brno, Czech Republic; Kardiovize study, International Clinical Research Centre, St. Anne’s University Hospital in Brno, Czech Republic; Department of Global Health and Population, Harvard TH Chan School of Public Health, Harvard University, USA; 2nd Department of Internal Medicine, St. Anne’s University Hospital in Brno and Faculty of Medicine, Masaryk University Brno, Czech Republic; Research Centre for Toxic Compounds in the Environment (RECETOX), Masaryk University, Czech Republic; Division of Epidemiology, Department of Quantitative Health Sciences, Mayo Clinic, USA; Division of Preventive Cardiology, Department of Cardiovascular Medicine, Mayo Clinic, USA; Marriot Heart Disease Research Program, Mayo Clinic, USA; Department of Neurosciences, Mayo Clinic, USA; Department of Neurology, and the Franke Global Neuroscience Education Center, Barrow Neurological Institute, USA; Division of Neurology, University Medical Centre, Slovenia; Celica Biomedical, Slovenia

**Keywords:** COVID-19, longitudinal, stress levels, depressive symptoms, risk factors

## Abstract

**Background:** Although several studies documented the impact of COVID-19 on mental health, the long-term effects of COVID-19 on mental health remain unclear.

**Aims:** To examine longitudinal changes in mental health prior to and during the consecutive COVID-19 waves in a well-established probability sample.

**Method:** An online survey was completed by the participants of the COVID-19 add-on study at 4 timepoints (N_1_=1823, N_2_=788, N_3_=532, N_4_=383): pre-COVID period (2014/2015), 1^st^ COVID-19 wave (April-May, 2020), 2^nd^ COVID-19 wave (August-October, 2020) and 3^rd^ COVID-19 wave (March-April, 2021). Data were collected via a set of validated instruments and analysed using latent growth models.

**Results:** During the pandemic, we observed a significant increase in stress levels (slope=1.127, P<0.001) and depressive symptoms (slope=1.177, P<0.001). The rate of increase in stress levels (cov=2.167, P=0.002), but not in depressive symptoms (cov=0.558, P=0.10), was associated with the pre-pandemic mental health status of the participants. Further analysis revealed two opposing clusters of factors that influenced mental health: loneliness and COVID-19 showed a negative effect on emotionality, while higher resilience acted protectively. A greater increase in stress was observed in women and younger participants.

**Conclusions:** The surge in stress levels and depressive symptoms persisted across all three consecutive COVID-19 waves. This surge is attributable to the effect of several risk factors including the status of mental health prior to the COVID-19 pandemic. Our findings have implications for strategies promoting resilience and addressing loneliness to mitigate the mental health impact of COVID-19 pandemic.

## Introduction

Several studies have reported changes in mental health immediately following the onset of the COVID-19 pandemic^1,2^. To date, however, only a limited number of studies have addressed the long-term effects of consecutive COVID-19 waves on mental health^3–11^. Some of these studies suggest that COVID-19 acts as a chronic stressor by direct impacts of the SARS-CoV-2 infection as well as indirectly by changing socio-economic, lifestyle and other circumstances of those experiencing the pandemic for longer than a year^9^.

In the Czech Republic, the 1^st^ COVID-19 wave spanned from mid-March to the end of May 2020 and coincided with the national lockdown with school closure and people instructed to stay at home (Figure 1). A relatively relaxed summer was then followed by an increase in SARS-Cov-2 infections and gradual resumption of restrictions. This 2^nd^ wave began in mid-August and lasted until December 2020. The 3^rd^ wave coincided with increased COVID-19-related hospitalization and mortality rates and the re-introduction of strict measures of the national lockdown, which started in January and lasted until the end of May 2021. While previous work documented significant surge in mental distress during the 1^st^ COVID-19 wave^12^, the long-term effects of the consecutive COVID-19 waves on the mental health in the Czech Republic remain unclear.

**Figure 1.**
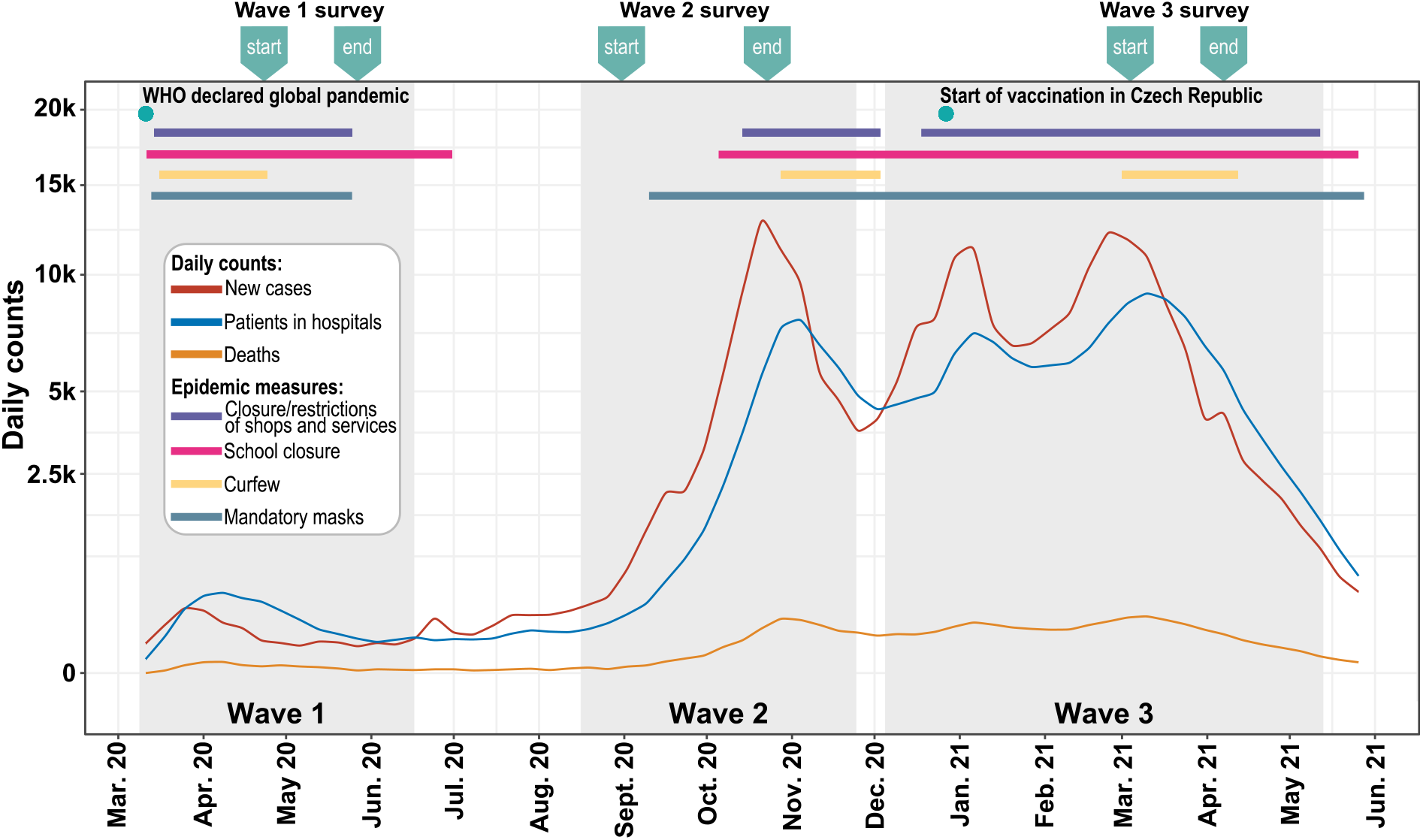
Time-line of the COVID-19 pandemic including restrictive measures in the Czech Republic. The lines show trends in daily numbers of new SARS-CoV-19 cases, SARS-CoV-19 related hospitalizations and deaths over the examined time period of the pandemic (data are projected on a square-root transformed scale). The upper horizontal bars show the time periods when key epidemiological measures were imposed. The grey sections indicate individual waves of the COVID-19 pandemic. The indicators above the plot show the time periods (start and end) of data collection in each COVID-19 wave. Source of the data: Ministry of Health of the Czech Republic (https://onemocneni-aktualne.mzcr.cz/covid-19).

Recent studies of the long-term effects of the COVID-19 pandemic on mental health reported significantly higher levels of stress^13,14^, anxiety^4,7^ and depressive symptoms^4,7,11^ during COVID-19 compared with the pre-COVID-19 period. Furthermore, younger age^9,15^, female gender ^4,16^, history of prior psychological distress^17^, worries and negative emotions about COVID-19^12,15,18,19^ and feeling of loneliness^9,13,16^ were identified as putative risk factors for developing mental health concerns during the COVID-19 pandemic. Some studies also suggested that adverse mental health outcomes reported during the COVID-19 may be long-lasting since they have been noted in people more than one year following SARS-CoV-2 infection^20^. Collectively, these observations are therefore suggestive of consecutive COVID-19 waves contributing to the long-term adverse mental health outcomes globally as well as in the Czech Republic. We here test this hypothesis by examining mental health prior to and across the consecutive COVID-19 waves in a probability population-based sample derived from the Czech Republic.

## Methods

### Study design and study population

This is a single-centre longitudinal COVID-19 ancillary study performed using the well-established Kardiovize population-based sample representing 1% of the inhabitants of the city of Brno, Czech Republic^12,21^. Participants of the Kardiovize study were selected randomly from the database of the health insurance companies to represent a probabilistic sample of the general population. The inclusion criteria for the COVID-19 add-on study were all participants of the Kardiovize study with available baseline pre-COVID-19 data on stress and depressive symptoms. Detailed descriptions of the recruitment processes for the Kardiovize and the COVID-19 add-on studies have been published elsewhere^21^. The COVID-19 add-on study followed the Strengthening the Reporting of Observational Studies in Epidemiology (STROBE) reporting guideline.

### Procedure

In the COVID-19 add-on study, we examined mental health in the same population-based sample prior to the COVID-19 pandemic and during the three consecutive COVID-19 waves (Figure 1). Mental health data were collected: 1) prior to the COVID-19 pandemic in 2014 and 2015, 2) during the 1^st^ COVID-19 wave, between April 23 and May 27 2020, 3) during the 2^nd^ COVID-19 wave, between August 31 and October 23 2020 and 4) during the 3^rd^ COVID-19 wave, between March 4 and April 7 2021. All participants completed the Kardiovize COVID-19 e-questionnaire (in Supplementary Appendix) including a battery of psychological scales through an online survey using validated RedCap software.

The primary outcomes were the stress levels, measured using the Perceived Stress Scale(PSS)^22^, and the severity of depressive symptoms, measured using the Patient Health Questionnaire(PHQ)^23^. PSS is a 10-item scale using 5-point Likert scale (0-Never to 5-Very often) with possible range of 0-40. Stress levels were categorized as low (score of 0-13), medium (score of 14-26) or high (score of 27-40). The reliability of the Czech version of the PSS in a single-factor approach was very good with ɑ=0.91^24^. Presence and severity of depressive symptoms were assessed using the identical two items from the PHQ-9 (prior to COVID-19) and the PHQ-4 (during COVID-19). Responses were measured using a 4-point Likert scale (0-Not at all to 3-Almost every day) with a possible range of 0-6 points. Despite its brevity, the reliability of the depression subscale proved to be good, ɑ=0.78^23^. Depressive symptoms were considered present if the sum of the score of the two PHQ items was equal to or greater than 3. When we talk collectively about moderate to high stress and increased depressive symptoms, we refer to them as mental health difficulties.

The secondary outcomes were related to psychosocial factors and different lifestyles. Perception of COVID-19 was assessed using the Brief-Illness Perception Questionnaire^25^. This 8-item scale uses a 10-point Likert scale (1-10) with possible range of 10-70 measures of the perception of COVID-19 illness as threatening across different domains. A meta-analytic study confirmed its good validity and reliability^26^. A two-factor approach assessing cognitive and emotional perceptions of COVID-19 illness was used. A higher score implies that COVID-19 illness is perceived as more threatening. The feeling of loneliness was assessed using the UCLA 3-item Loneliness Scale^27^. A 3-point Likert scale (1-Seldom to 3-Often) providing a total score in the range of 3-9 points showed good reliability of ɑ=0.89-0.94^28^. Higher scores indicate greater feelings of loneliness. Presence of the feelings of loneliness was defined as UCLA score equal to or greater than 6. Resilience was assessed using the Connor-Davidson Resilience Scale^29^. This short 2-item instrument with a 5-point Likert scale (0-Not at all to 4-Almost always) and a score range of 0-8 demonstrated very good validity and reliability with ɑ=0.79^29,30^. Presence of resilience was defined as low (score of 0-5), medium (score of 6-7) or high (score of 8). Resilient coping was assessed using the Brief Resilient Coping Scale^31^. It is a 4-item scale using a 5-point Likert scale (1-5) with a possible range of 4-20 points. Previous studies have confirmed good validity and reliability of the BRCS with ɑ=0.78^32^. Higher score represents better (more positive) way of coping with difficult situations with the low (score of 4-13), medium (score of 14-16) and high (score of 17-20) categories.

All participants also provided general demographic and health data and answered several questions about how COVID-19 lockdown affected their lifestyle and in general about their experience with the COVID-19 lockdown as part of the Kardiovize COVID-19 e-questionnaire (in Supplementary Appendix).

### Statistical analysis

Missing values correspond only to respondents who dropped out in each wave. No imputation of missing values was applied. The Wilcoxon rank-sum test, and chi-square test were used to verify representativeness of COVID-19 study population. Differences in the prevalence of mental health difficulties were verified using the G-test with Benjamini-Hochberg correction for multiple comparisons. Longitudinal changes in the severity of depressive symptoms and stress levels and the effect of risk factors on these changes were analysed using latent growth curve models (LGCM). Given the non-normal distribution of some variables, we have used and report a robust estimator (maximum likelihood estimation with robust (Huber-White) standard errors). All 2-sided P<0.05 were considered to be significant. Data analysis was performed in the RStudio (v.2022.02.3, R version 4.2.0).

### Ethics statement

The authors assert that all procedures contributing to this work comply with the ethical standards of the relevant national and institutional committees on human experimentation and with the Helsinki Declaration of 1975, as revised in 2008. All procedures involving human subjects/patients were approved by the Internal Review Board and the St. Anne’s University Hospital ethics committee. All adult participants provided written informed consent to participate in this study.

## Results

### Sample characteristics and representativeness

A total of 1823 Kardiovize study participants completed baseline pre-COVID-19 assessment including stress levels (PSS) and severity of depressive symptoms (PHQ) scales. These Kardiovize participants were electronically invited to join the COVID-19 add-on study. A total of 43% (N=788) of the Kardiovize participants enrolled in the COVID-19 add-on study and completed the survey in at least one wave. The COVID-19 add-on study sample in general maintained the representativeness of the original Kardiovize study population. Specifically, sex (P=0.98), education (P=0.99), job position (P=0.99), pre-pandemic stress levels (P=0.50) and severity of depressive symptoms (P=0.44) were comparable between the Kardiovize and the COVID-19 add-on study cohorts. Only age differed between the two groups with a difference in mean age of 1.32 years (mean[95%CI], Kardiovize population, 47.3[46.83, 47.78], COVID-19 population, 45.98[45.17, 46.78], P=0.004). The online survey was completed during the 2^nd^ and 3^rd^ COVID-19 waves by 67.5% (N=532) and 48.6% (N=383) of the COVID-19 add-on study participants. The basic demographic characteristics are shown in Table 1.

**Table 1.**
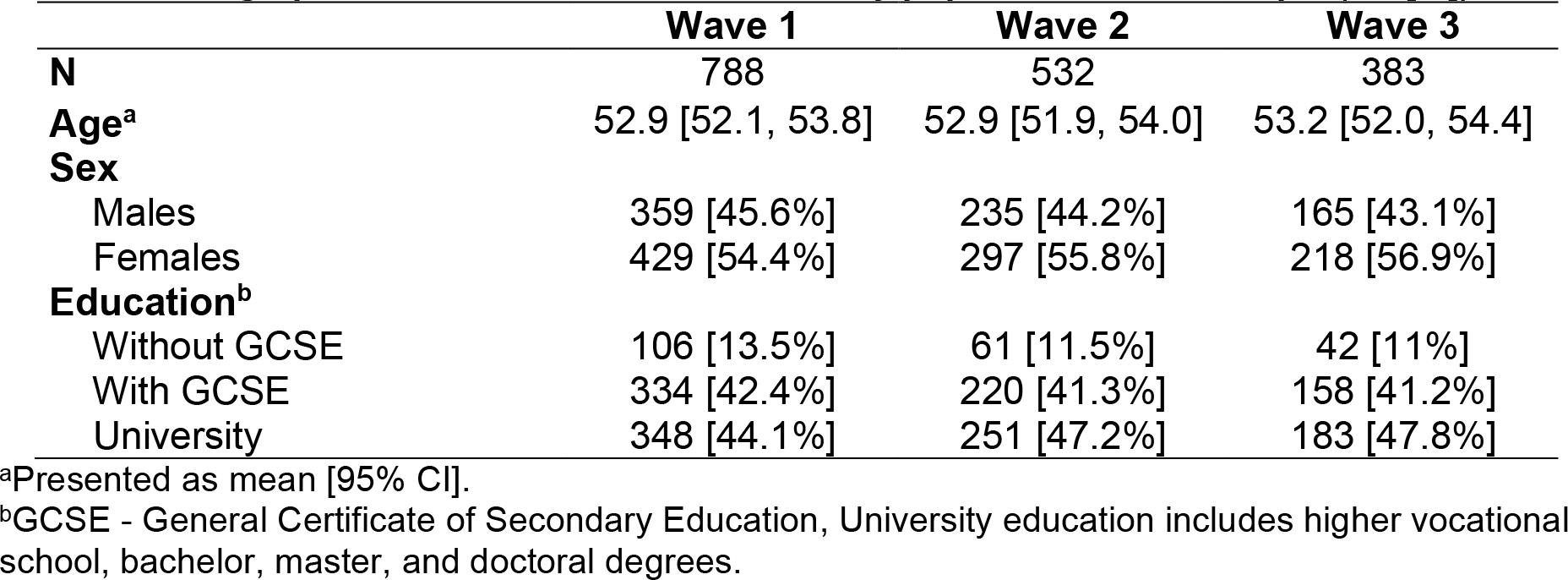
Demographics of the COVID-19 add-on study population-based sample (No [%]).

|  | **Wave 1** | **Wave 2** | **Wave 3** |
| --- | --- | --- | --- |
| **N** | 788 | 532 | 383 |
| **Age<sup>a</sup>** | 52.9 [52.1, 53.8] | 52.9 [51.9, 54.0] | 53.2 [52.0, 54.4] |
| **Sex** |  |  |  |
| Males | 359 [45.6%] | 235 [44.2%] | 165 [43.1%] |
| Females | 429 [54.4%] | 297 [55.8%] | 218 [56.9%] |
| **Education<sup>b</sup>** |  |  |  |
| Without GCSE | 106 [13.5%] | 61 [11.5%] | 42 [11%] |
| With GCSE | 334 [42.4%] | 220 [41.3%] | 158 [41.2%] |
| University | 348 [44.1%] | 251 [47.2%] | 183 [47.8%] |

<sup>a</sup>Presented as mean [95% CI].
<sup>b</sup>GCSE - General Certificate of Secondary Education, University education includes higher vocational school, bachelor, master, and doctoral degrees.

### Changes in stress levels

The mean stress levels (mean[95% CI]) increased during the 1^st^ (13.8[13.3, 14.3]), 2^nd^ (12.9[12.1, 13.3]) and the 3^rd^ COVID-19 waves (13.9[13.1, 14.6]) compared with the pre-COVID-19 period (12.0[11.6, 12.5]) (Figure 2A). The prevalence of moderate-to-high stress also increased from 44.6% in the pre-COVID-19 period to 49.5% and 51.4% during the 1^st^ and 3^rd^ COVID-19 waves, respectively, and was lower (43.2%) in the 2^nd^ wave (Figure 2B). The prevalence of moderate-to-high stress in the consecutive waves was on average 1.2, 1.11, and 1.22 times higher in women compared with men. The growth model (Table 2.A) confirmed that stress levels increased significantly over time (std. β_slope_=1.127, P<0.001). Individual respondents also differed in baseline stress level (std. β_intercept_=3.735, P<0.001). The interaction of slope and intercept showed that those with higher baseline stress exhibited further increase in stress levels over time, while those with lower baseline stress tended to decrease stress levels in due course of the pandemic (std. cov(s,i)=2.167, P=0.002, Figure 2C).

**Table 2.** LGCM goodness-of-fit indices and main estimates for severity of depressive symptoms and stress level.

| **A. Stress level** |  |  |  | **B. Severity of depressive symptoms** |  |  |
| --- | --- | --- | --- | --- | --- | --- |
|  | **Value** |  |  | **Value** |  |  |
| X<sup>2</sup> [DF] | 10.267 [3] |  |  | 10.098 [3] |  |  |
| X<sup>2</sup> P-value | 0.016 |  |  | 0.018 |  |  |
| CFI | 0.988 |  |  | 0.976 |  |  |
| TLI | 0.976 |  |  | 0.952 |  |  |
| RMSEA | 0.085 |  |  | 0.079 |  |  |
|  | **Estimate[SE]** | **Std. $\beta$** | **P** | **Estimate[SE]** | **Std. $\beta$** | **P** |
| Slope (s) | 2.279[0.397] | 1.127 | <0.001 | 0.850[0.076] | 1.177 | <0.001 |
| Intercept (i) | 11.5[0.317] | 3.735 | <0.001 | 0.588[0.054] | 1.034 | <0.001 |
| Covariance (s,i) | 13.487[4.355] | 2.167 | 0.002 | 0.229[0.139] | 0.558 | 0.1 |

**Figure 2.**
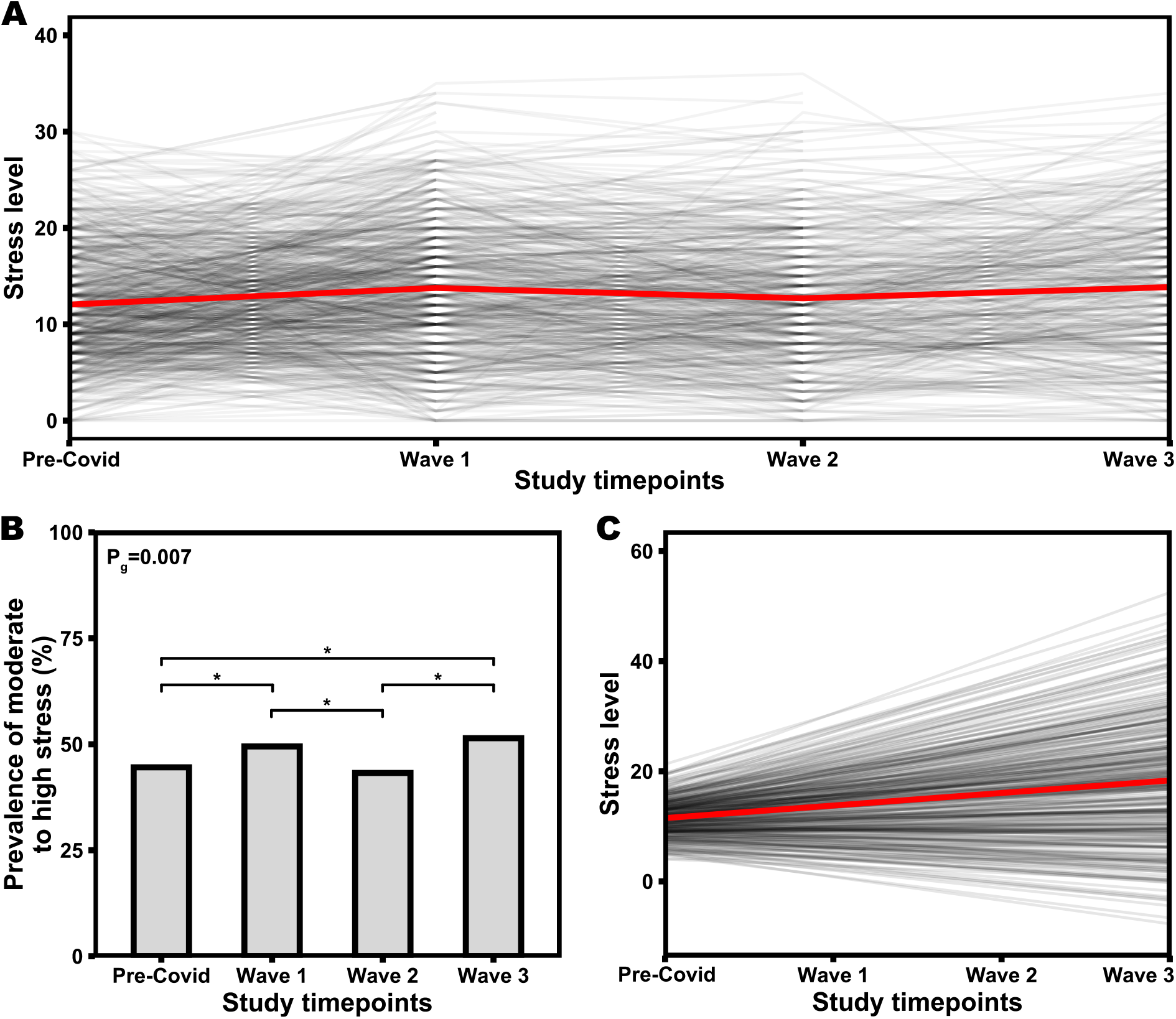
Stress levels prior to and during the consecutive COVID-19 waves. The lines show the changes in the perceived stress scores at each time point for individual participants; the red line indicates the evolution of the mean score. B) Bar plot depicts the prevalence of moderate-to-high stress (PSS score ≥ 14) prior to and during the three COVID-19 waves. Upper horizontal bars denote significant differences between time points (*P<0.05). C) Lines show the predicted trend in stress levels during the pandemic for individual participants based on the latent growth curve model, the red line shows the average trend (slope).

### Changes in the severity of depressive symptoms

The severity of depressive symptoms (mean[95% CI]) increased during the 1^st^ (1.2[1.1, 1.3]), 2^nd^ (1.0[0.8, 1.1]) and the 3^rd^ COVID-19 waves (1.4[1.3, 1.6]) compared with the pre-COVID-19 period (0.7[0.6, 0.7]) (Figure 3A). Similarly, the prevalence of significant depressive symptoms increased from 13.3% in the pre-COVID-19 period to 16% and 20.4% during the 1^st^ and 3^rd^ COVID-19 waves, respectively, while it showed no changes (13.2%) during the 2^nd^ wave (Figure 3B). The prevalence of significant depressive symptoms in concomitant waves was on average 1.28, 1.11 and 1.31 times higher in women compared with men, respectively. The growth model (Table 2.B) confirmed that the severity of depressive symptoms increased significantly over time (std. β_slope_=1.177, P<0.001). Individual respondents also differed in baseline depressive symptom severity (std. β_intercept_=1.034, P<0.001), with those who had greater depressive symptoms at baseline showing a trend toward greater increases over the course of the pandemic (std. cov(s,i)=0.558, P=0.10, Figure 3C).

**Figure 3.**
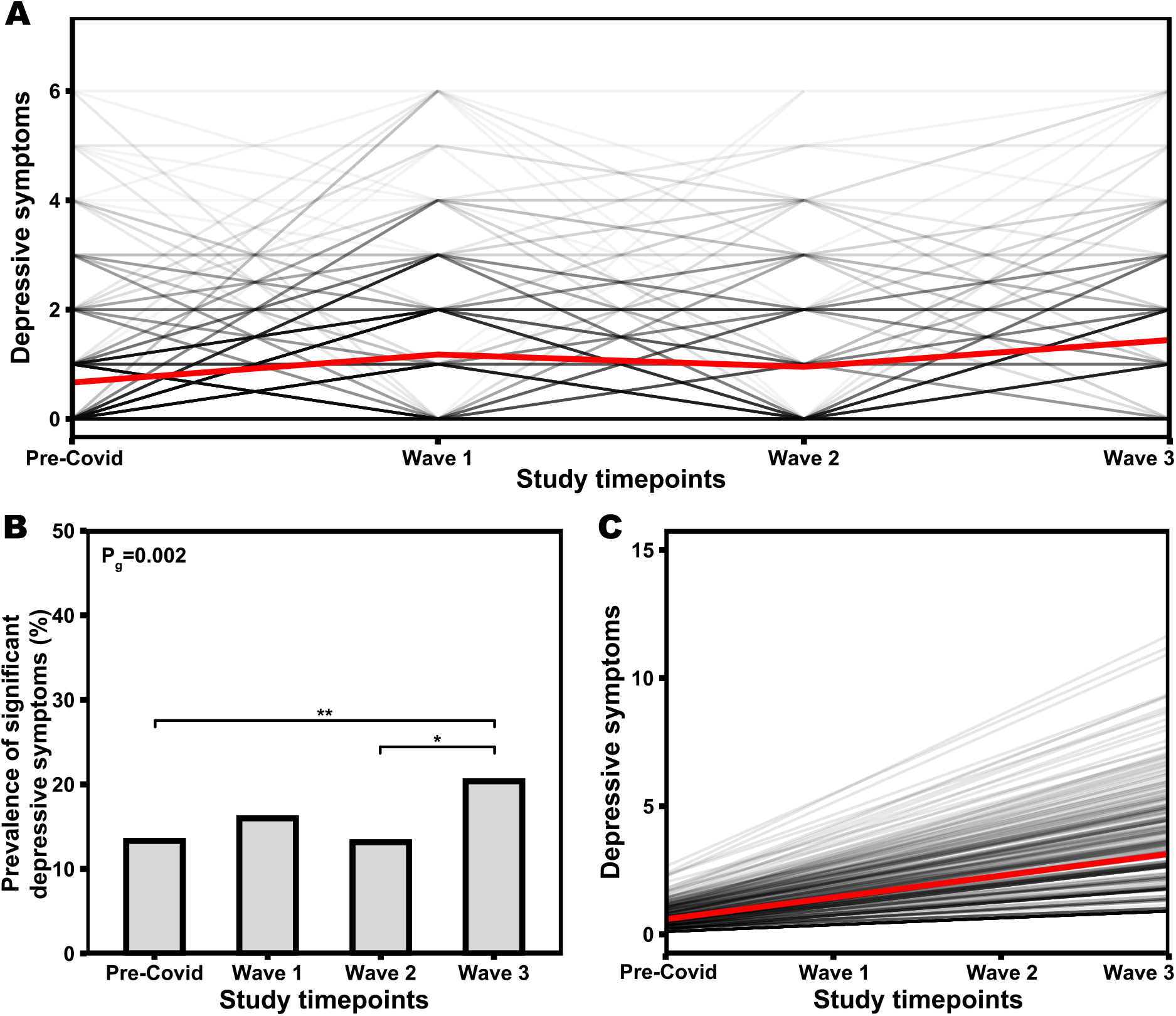
Severity of depressive symptoms prior to and during the consecutive COVID-19 waves. The lines show the changes in the severity of depressive symptoms at each time point for individual participants; the red line indicates the evolution of the mean score. B) Bar plot depicts the prevalence of significant depressive symptoms (PHQ score ≥ 3) prior to and during the three COVID-19 waves. Upper horizontal bars denote significant differences between time points (*P<0.05, **P<0.01). C) Lines show the predicted trend in severity of depressive symptoms during the pandemic for individual participants based on the latent growth curve model, the red line shows the average trend (slope).

### The effects of the risk factors

Finally, we investigated the potential effect of psychosocial variables and lifestyle factors on mental health difficulties during the COVID-19 pandemic. Results showed that feelings of loneliness, emotional perception of COVID-19 as threatening, and being older had the greatest effect on the increase in perceived stress. Inversely, higher levels of resilience served as a buffer against increased stress. Further, men had lower baseline stress levels, but the trajectory of stress levels over time did not differ between men and women. Lastly, we also observed a negative effect of a poor financial situation during the first wave and higher perceived stress among respondents who experienced COVID-19 during the second and third waves and who cognitively perceived COVID-19 as threatening (Figure 4A, Table 3.A). Intriguingly, when we controlled for the effect of risk factors, we did not observe any changes in stress levels over time (i.e., during the COVID-19 pandemic); on the contrary, there was a decrease in stress levels in this context. (Figure 3.B).

**Table 3.** LGCM goodness-of-fit and estimates of the effect of risk factors on longitudinal changes in the severity of depressive symptoms and stress level.

| A. Stress level |  |  |  | B. Severity of depressive symptoms |  |  |
| --- | --- | --- | --- | --- | --- | --- |
|  | Value |  |  | Value |  |  |
| X<sup>2</sup> [DF] | 93.953[37] |  |  | 47.866[25] |  |  |
| X<sup>2</sup> P-value | <0.001 |  |  | 0.004 |  |  |
| CFI | 0.938 |  |  | 0.946 |  |  |
| TLI | 0.9 |  |  | 0.916 |  |  |
| RMSEA | 0.067 |  |  | 0.053 |  |  |
|  | Estimate[SE] | Std. B | P | Estimate[SE] | Std. B | P |
| Slope (s) | -1.931[1.323] | -0.146 | 0.144 | 0.104[0.285] | 0.162 | 0.716 |
| Intercept (i) | 13.85[2.087] | 4.155 | <0.001 | 0.262[0.383] | 0.634 | 0.493 |
| Covariance (s,i) | 1.45[1.407] | 0.6 | 0.303 | 0.161[0.092] | 1.084 | 0.125 |
| *Time invariant risk factors* |  |  |  |  |  |  |
| Age~slope | 0.027[0.011] | 0.457 | 0.015 |  |  |  |
| Sex (male)~slope | 0.032[0.28] | 0.021 | 0.910 |  |  |  |
| Age~intercept | -0.05[0.02] | -0.195 | 0.015 |  |  |  |
| Sex (male)~intercept | -1.436[0.51] | -0.214 | 0.005 |  |  |  |
| *Time variant risk factors* |  |  |  |  |  |  |
| Finances (Wave1) | 1.096[0.321] | 0.123 | 0.001 | 0.239[0.082] | 0.133 | 0.003 |
| Loneliness (Wave1) | 0.678[0.151] | 0.171 | <0.001 | 0.202[0.038] | 0.254 | <0.001 |
| Resilience (Wave1) | -1.217[0.184] | -0.231 | <0.001 | -0.167[0.045] | -0.157 | <0.001 |
| Emot. COVID-19 perception (Wave1) | 0.510[0.07] | 0.338 | <0.001 | 0.075[0.013] | 0.248 | <0.001 |
| Cogn. COVID-19 perception (Wave1) | 0.061[0.036] | 0.056 | 0.089 |  |  |  |
| Finances (Wave2) | 0.323[0.32] | 0.036 | 0.314 | -0.001[0.083] | 0 | 0.992 |
| Loneliness (Wave2) | 0.948[0.158] | 0.225 | <0.001 | 0.203[0.041] | 0.245 | <0.001 |
| Resilience (Wave2) | -1.262[0.175] | -0.251 | <0.001 | -0.151[0.034] | -0.154 | <0.001 |
| Emot. COVID-19 perception (Wave2) | 0.331[0.065] | 0.222 | <0.001 | 0.087[0.014] | 0.299 | <0.001 |
| Cogn. COVID-19 perception (Wave2) | 0.142[0.041] | 0.131 | 0.001 |  |  |  |
| COVID-19 experience (Wave2) | 4.495[0.54] | 0.053 | <0.001 |  |  |  |
| Finances (Wave3) | 0.576[0.379] | 0.059 | 0.128 | 0.089[0.104] | 0.044 | 0.391 |
| Loneliness (Wave3) | 0.847[0.146] | 0.215 | <0.001 | 0.184[0.041] | 0.227 | <0.001 |
| Resilience (Wave3) | -1.380[0.214] | -0.228 | <0.001 | -0.161[0.051] | -0.129 | 0.001 |
| Emot. COVID-19 perception (Wave3) | 0.51[0.066] | 0.334 | <0.001 | 0.097[0.016] | 0.307 | <0.001 |
| Cogn. COVID-19 perception (Wave3) | 0.107[0.045] | 0.1 | 0.016 |  |  |  |
| COVID-19 experience (Wave3) | 1.422[0.608] | 0.081 | 0.019 |  |  |  |

**Figure 4.**
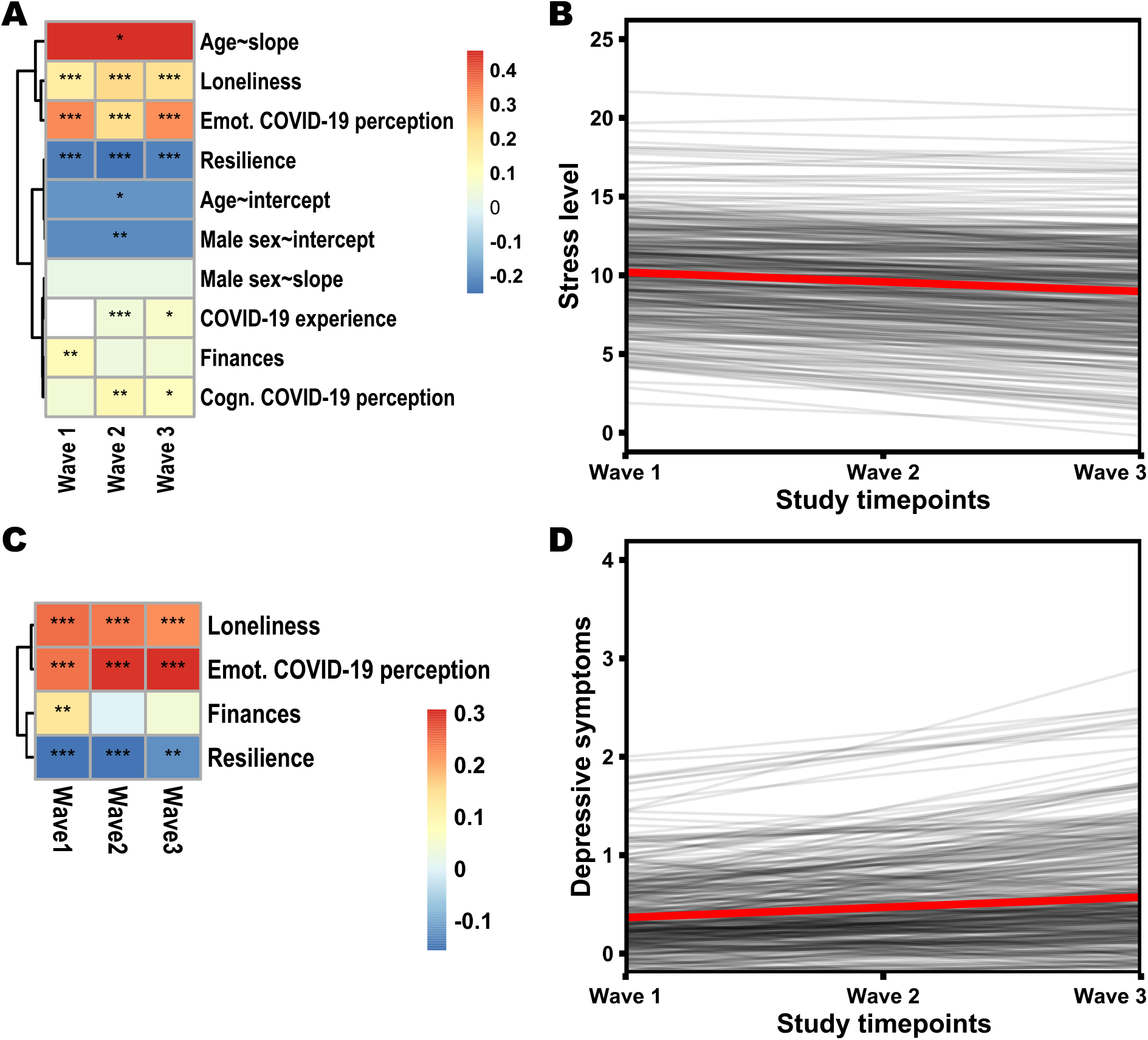
Effect of risk factors on longitudinal changes in stress level and severity of depressive symptoms during the consecutive COVID-19 waves. A) Heatmap shows standardized beta coefficients of the effect of time-variant and time-invariant risk factors on stress levels (only factors with a significant effect are included). Asterisks indicate the level of significance (*P<0.05, **P<0.01, ***P<0.001). B) Lines show the predicted trend in stress levels over the course of the pandemic for individual participants based on the latent growth curve model when controlling for the influence of significant risk factors (A), the red line shows the average trend (slope). C) Heatmap shows standardized beta coefficients of the effect of time-variant risk factors on severity of depressive symptoms (only factors with a significant effect are included). Asterisks indicate the level of significance (**P<0.01, ***P<0.001). D) Lines show the predicted trend in severity of depressive symptoms over the course of the pandemic for individual participants based on the latent growth curve model when controlling for the influence of significant risk factors (C), the red line shows the average trend (slope).

The severity of depressive symptoms was affected by fewer risk factors. Unlike stress, we did not observe an effect of sex or age. We again observed a negative effect of feelings of loneliness and negative emotional perception of the COVID-19 illness throughout the pandemic, and higher depressive symptoms due to worse financial situation in the first wave. Furthermore, for depressive symptoms, we demonstrate a protective effect of higher resilience (Figure 3C, Table 3.B). Analogous to stress, when we controlled for the effect of risk factors, the levels of depressive symptoms did not change significantly over time (Figure 4D).

## Discussion

Stress levels and severity of depressive symptoms demonstrated significant increase during the COVID-19 pandemic, and the prevalence of increased stress and significant depressive symptoms was greater during the pandemic compared to pre-pandemic. Self-reported repeated testing of parameters indicative of mental health difficulties prior to and across the COVID-19 waves in the Czech Republic, therefore, provided results consistent with several similar longitudinal studies reported recently from other countries including the UK^9,14^, Germany^33^, Canada^11^, Switzerland^34^, and France^4^, a recent meta-analytical study^35^ as well as numerous cross-sectional studies^36–38^. These studies, altogether, embrace the hypothesis that consecutive COVID-19 waves produced long-term deterioration of mental health.

Furthermore, the results indicate that the trend in mental health difficulties depended on the pre-pandemic levels of stress and depressive symptoms. Higher pre-pandemic levels of stress and depressive symptoms were associated with a greater growth in these difficulties during the pandemic. For respondents who had low pre-pandemic levels of mental health difficulties, the severity of depressive symptoms increased only minimally during the pandemic, and stress levels even decreased in these individuals with time. Differences in individual trajectories suggest that the ability to cope with the burden imposed by the COVID-19 pandemic is to some extent dependent on the overall level of coping resources available to the individual and their durability or rate of depletion in coping with the pandemic^8^.

One of the key questions, the answer to which is essential for the formulation of adequate help and intervention, are the mechanisms underlying these changes. To uncover these mechanisms, we investigated the effects of a series of potential risk factors on the changes in mental health difficulties during the COVID-19 pandemic.

The investigation identified a cluster of factors that affected stress levels and severity of depressive symptoms. The risk factors that negatively contributed to both of these indicators of mental health difficulties were associated with the emotional experience of the circumstances of the COVID-19 pandemic, namely feelings of loneliness and the negative effect of COVID-19 disease on emotions. These results support previous findings of a significant link between negative emotionality and an increase in mental health difficulties ^9,12,15,16,18,39,40^. Resilience, on the other hand, demonstrated a protective, buffering effect, consistent with its very definition, the ability to overcome adversity without compromising its own mental and emotional stability^36,41–43^.

Besides these two main clusters of influences, we also observed the effects of age and sex on the stress levels. Women and younger respondents showed a greater surge in stress compared to men. The sex-related differences are consistent with previous studies^44–46^. Possible reasons for this finding consist of stronger emotional experiencing of women or the greater impact of daily life limitations due to pandemic measures in difficulties in balancing family and work^47^. Greater levels of stress in younger participants are likely to be associated with disrupted social interactions, greater worries about studies, job security and financial stability, and richer life experiences and reduced life expectations in the older ones^35,48,49^.

Intriguingly, when we included the effects of risk factors into the model, the trend in mental health difficulties was no longer significantly increasing with time. The average severity of depressive symptoms now remained constant, while we could even observe a slight decrease in stress levels. This hints that the surge in mental health difficulties observed globally in relation to the COVID-19 pandemic is attributable particularly to the negative (or, conversely, protective) effects of these key risk factors. Given the smaller cohort size, however, further studies are needed to confirm this exceptional and significant role of the risk factors. We also observed differences in the response to COVID-19 based on the pre-pandemic levels of stress and depressive symptoms. Among those who had already higher levels of depressive symptoms before the pandemic, we observed further increase during the pandemic. Similarly for stress, the rate of decline appeared to be smaller for respondents with higher pre-pandemic levels than for others. It appears that, especially in the case of depressive symptoms, additional negative circumstances and adversities (beyond the described risk factors) are likely to affect the emotional state of individuals.

### Strengths and limitations

This study has several strengths. First, we have described changes in mental health difficulties in several distinctive waves of the COVID-19 pandemic that differed in their characteristics, severity, related measures, duration, and thereby impact on people’s daily lives. In addition, we compared these changes with the situation prior to the outbreak of the COVID-19 pandemic. Secondly, and most importantly, in contrast to most other large population surveys and other research studies, we analysed the impact of multiple key risk factors at once, measured using standardised psychological instruments. Compared with many other studies that have included only the influence of basic socio-demographic characteristics or one or two risk factors, we can thereby portray a more comprehensive picture of the longitudinal evolution of mental health during a pandemic and therefore outline areas that require further research in larger or socially and culturally diverse populations.

There are also several limitations that should be mentioned. Although the observations about mental health prior to and during the three consecutive COVID-19 waves reported in this study derive from a probability population-based sample, the examined sample size of 788 participants is relatively small compared with national surveys^7^ and some other studies^9,33^. Although this is a representative population sample, the smaller number of respondents may affect the generalisability of the described findings, which would benefit from further confirmation on larger samples. The drop-out rates recorded in our study, however, are lower than the ones reported in other longitudinal studies of mental health prior and/or during the COVID-19 waves^9,33,50^. Similarly to other studies^9,50^, we also used self-reported psychological scales and questionnaires to estimate changes in mental health in response to consecutive COVID-19 waves. Therefore, our results are based on the examination of symptoms rather than formal clinical diagnoses of mental health disorders. As COVID-19 add-on cohort was sampled exclusively during the COVID-19 waves, our study precludes testing whether mental distress observed during the COVID-19 waves subsided in the time intervals between the COVID-19 waves.

## Conclusions and recommendations

We here show that increased stress levels and severity of depressive symptoms observed at the onset of the COVID-19 pandemic in comparison with the pre-COVID-19 period persist throughout all of the consecutive COVID-19 waves. This finding is indicative of long-term effects of COVID-19 on mental health and implicates several direct and indirect stressors, which will likely also play a critical role in determining the time needed for the mental health difficulties to eventually subside following the end of the COVID-19 pandemic. Incorporation of more efficient and targeted approaches to mental health care, more timely strategies to identify and treat individuals at high risk for developing mental health difficulties^51,52^ and proactive use of online technologies and web-based intervention tools^53^ could help curb mental health difficulties that arose during the COVID-19 pandemic.

## Data Availability

The data used in this study are available on request immediately following the publication to anyone who submits the online request that will be approved by the St. Anne's University Hospital International Clinical Research Centre internal board. The researches have to provide their research intentions and goals, and specify, and justify requested variables. The data will be provided for a limited and well-defined time via cloud service or e-mail in csv format. After defined period the data should be returned, and all other copies should be destroyed.

## Statements

### Author Contribution

The authors contributed to this article as follows: GBS and JPGR conceived the idea of this study, SK, MS, JSN, APos, and APol contributed to the discussion and battery creation, JSN and Apos verified the data, JSN made the statistical analysis, JSN and GBS wrote the draft of the manuscript, MS, YEG, JRMI, FLJ, MV, GBS contributed to the discussion and critical review of the manuscript.

### Data Availability

The data that support the findings of this study are available from the corresponding author, GBS, upon substantiated request and approval by the St. Anne’s University Hospital International Clinical Research Centre internal board.

## Supplementary Appendix

### Kardiovize COVID-19 e-questionnaire (excluding standardized questionnaires)

*If the question was used only in one of the data collections or has been modified, this is indicated in italics in square brackets after the question text)*

1. What is your current weight? *[Data collection 1]*
2. How many cigarettes do you smoke per day? If you are a non-smoker, please, fill 0. *[Data collection 1]*
3. What is your current family situation?
  a. Living in the relationship with the children
  b. Living in the relationship without the children
  c. Monoparental household (living with one parent)
  d. Living alone
  e. Other (please, specify)
4. How many children do you have? *[Data collection 1]*
  a. None
  b. One child
  c. Two children
  d. Three or more children
5. Who are you spending your time with during the quarantine? (multiple choice)
  a. No one
  b. With my partner or spouse
  c. With my children
  d. With other family members
  e. With someone outside my own family
6. During the last 14 days, how often have you been actively and specifically seeking information about the current situation regarding the COVID-19 pandemic and related measures?
  a. Never
  b. Less than once per week
  c. 1-2 times per week
  d. 2-3 times per week
  e. Approximately once per day
  f. Many times per day
7. Does the COVID-19 state of emergency affect your financial situation?
  a. Not at all
  b. Just a little
  c. Pretty much
  d. Extremely
8. How much does the current COVID-19 situation affect your work life? (multiple choice)
  a. The pandemic did not affect my work life/I am currently not working
  b. I have more work than usual
  c. I have less work than usual
  d. I work from home
  e. I changed my job/my job duties or position changed
  f. I stayed home because of kids or family member
  g. I lost my job
9. How many individual PRIVATE (not work-related) social contacts (phone, SMS, Skype, WhatsApp, email, …) have you had in the last 7 days? *[Data collection 1]*
  a. None, I am without social contacts
  b. 1 to 3 contacts
  c. 4 - 7 contacts
  d. 8 - 14 contacts
  e. 15 and more contacts
10. How many individual WORK-RELATED social contacts (phone, SMS, Skype, WhatsApp, email, …) have you had in the last 7 days? *[Data collection 1]*
  a. None
  b. 1 to 3 contacts
  c. 4 - 7 contacts
  d. 8 - 14 contacts
  e. 15 and more contacts
11. Has your sleep quality changed in the past last 14 days/compared to last time you filled this survey?
  a. it got better
  b. it not changed
  c. it got worse
12. Has the length of your sleep changed (on average per day) /compared to last time you filled this survey?
  a. sleep time has increased
  b. sleep time did not change
  c. sleep time has decreased
  How often have you exercised in the last 14 days? Write down how many hours per week have you spent performing specific exercises, if zero time, fill in 0.
13. Low intensity exercise (e.g. walking):
14. High intensity exercise (e.g. running):
15. Body building:
16. Stretching:
17. Has the frequency of how often you exercise changed over the last 14 days/compared to last time you filled this survey?
  a. the frequency has increased
  b. the frequency has not changed
  c. the frequency has decreased
18. How many times per week did you go out from your home (work, shop, nature, etc.) in the last 14 days?
  a. Never
  b. 1-2 times per week
  c. 3-5 times per week
  d. Almost every day
  **How do you follow the government-imposed COVID-19 state of emergency measures?**
19. Are you wearing a mask/respirator?
  a. Always
  b. Almost always
  c. Sometimes
  d. Never
20. How often are you washing or disinfecting your hands?
  a. Always
  b. Almost always
  c. Sometimes
  d. Never
21. How often have you respected the restriction of going out?
  a. Always
  b. Almost always
  c. Sometimes
  d. Never
22. How often have you respected the 2-meter social distancing?
  a. Always
  b. Almost always
  c. Sometimes
  d. Never
23. How often have you respected the ban of direct contact with other people?
  a. Always
  b. Almost always
  c. Sometimes
  d. Never
24. How often have you respected the measure that only two people can be in closer contact in public places?
  a. Always
  b. Almost always
  c. Sometimes
  d. Never
25. Do you trust the government and other institutions involved in how they are dealing with the current situation? [*Data collections 2 and 3]* 1 = Do not trust at all - 5 = Absolutely trust
26. Are current anti-epidemic measures adequate? [*Data collections 2 and 3]* 1 = Completely inadequate - 5 = Completely adequate
27. When do you think the life will get back to the normal in the Czech Republic*? Please indicate the number of months*.
28. How many days did you spend in isolation? *If you have not been in quarantine, please fill in 0*.
  a. because of contact with a person with confirmed coronavirus infection:
  b. because returning from a COVID-19 high risk country:
  c. because you have been tested positive for coronavirus infection:
29. Have you been vaccinated with the Covid-19 vaccine? *[Data collection 3]*
  a. Yes
  b. No
30. What is your willingness to be vaccinated with the Covid-19 vaccine? *[Data collection 3]* 1 = Refuse to be vaccinated - 5 = Definitely want to be vaccinated
31. Have you fell ill with COVID-19?
  a. Yes
  b. No
32. What symptoms or signs of COVID-19 have you manifested? (multiple choice)
  c. Fever
  d. Runny nose and cough
  e. Emphysema
  f. Pneumonia
  g. Loss of taste and smell
  h. Headache and dizziness
  i. Nausea, vomiting, diarrhea
  j. Weakness, joints and muscle pain
  k. No symptoms or signs
  **In the following section we will ask you about your health**. *[Data collection 1]*
33. Have you been treated for arterial hypertension?
  a. Yes
  b. No
34. If yes, please provide the name of the medication and the dosage to treat arterial hypertension:
35. Have you been diagnosed with diabetes mellitus type I?
  a. Yes
  b. No
36. If yes, please provide the name of the medication and the dosage to treat diabetes mellitus type I:
37. Have you been diagnosed for diabetes mellitus type II?
  a. Yes
  b. No
38. If yes, please provide the name of the medication and the dosage to treat diabetes mellitus type II:
39. Have you been diagnosed with disease of respiratory-tract (astma bronchiale, CHOPN, etc.)?
  a. Yes
  b. No
40. If yes, please provide the name of the medication and the dosage to treat the respiratory-track disease:
41. Have you been diagnosed with any of the following immune disorders?
  a. Inflammatory bowel disease (e.g. ulcerative colitis or Crohn’s disease)
  b. Rheumatic diseases (e.g. rheumatoid arthritis)
  c. Multiple sclerosis (MS)
  d. Bone marrow transplant
  e. Organ transplant and immunosuppressive therapy
  f. Cancer treated with chemotherapy or radiotherapy
  g. None
42. If yes, please provide the name of the medication and the dosage to treat these disorders:
43. Have you been diagnosed with an allergy or atopic eczema?
  a. Yes
  b. No
44. Are you currently taking medicines containing corticosteroid (e.g. Decamed, Medrol, Depo-Medrol, Dexamethasone, Hydrocortisone, Fortecortin, Methycetone, Fludrocortisone)?
  a. Yes
  b. No
45. Are you currently taking medicines containing Hydrochloroquine (e.g. Plaquenil)?
  a. Yes
  b. No

